# Ignoring transmission dynamics leads to underestimation of the impact of a novel intervention against mosquito-borne disease

**DOI:** 10.1101/2021.11.19.21266602

**Authors:** Sean M. Cavany, John H. Huber, Annaliese Wieler, Quan Minh Tran, Manar Alkuzweny, Margaret Elliott, Guido España, Sean M. Moore, T. Alex Perkins

**Author notes:** For correspondence: Sean Cavany, John Huber, and Alex Perkins. Contributed Equally (Sean M. Cavany, John H. Huber).

## Abstract

New vector-control technologies to fight mosquito-borne diseases are urgently needed, the adoption of which depends on efficacy estimates from large-scale cluster-randomized trials (CRTs). The release of *Wolbachia*-infected mosquitoes is one promising strategy to curb dengue virus (DENV) transmission, and a recent CRT reported impressive reductions in dengue incidence following the release of these mosquitoes. Such trials can be affected by multiple sources of bias, however. We used mathematical models of DENV transmission during a CRT of *Wolbachia*-infected mosquitoes to explore three such biases: human movement, mosquito movement, and coupled transmission dynamics between trial arms. We show that failure to account for each of these biases would lead to underestimated efficacy, and that the majority of this underestimation is due to a heretofore unrecognized bias caused by transmission coupling. Taken together, our findings suggest that *Wolbachia*-infected mosquitoes could be even more promising than the recent CRT suggested. By emphasizing the importance of accounting for transmission coupling between arms, which requires a mathematical model, our results highlight the key role that models can play in interpreting and extrapolating the results from trials of vector control interventions.

## Introduction

Dengue virus (DENV) poses a risk to around half the world’s population due to the widespread abundance of its *Aedes* mosquito vectors [1]. Historically, the success of dengue control has been limited by challenges such as the expanding distribution of *Aedes aegypti* due to urbanization and land-use changes, and ineffective or sub-optimally applied control strategies [2,3]. One novel control strategy that holds promise is the release of mosquitoes infected with *Wolbachia*, a vertically transmitted intracellular bacteria that reduces the ability of *Aedes aegypti* mosquitoes to transmit DENV [4]. A cluster-randomized, controlled trial conducted between 2018 and 2020 in Yogyakarta, Indonesia (Applying Wolbachia to Eliminate Dengue, AWED) [5,6] estimated that release of *Wolbachia*-infected mosquitoes had a protective efficacy against symptomatic, virologically confirmed dengue of 77.1% (95% confidence interval: 65.3-84.9%) [7].

There are at least three factors that can result in underestimated efficacy in this type of trial. All operate by making outcomes in treatment and control clusters appear more similar than if these factors were not at play, although they result in this for different reasons. First, the movement of humans between control and treated clusters can increase the exposure to DENV of study subjects residing in treatment clusters and lower the exposure of subjects residing in control clusters [8]. Second, the movement of mosquitoes between arms can lead to an appreciable proportion of mosquitoes in control clusters infected with *Wolbachia*, lowering these mosquitoes’ ability to transmit DENV and introducing a source of contamination across trial arms. Third, the dynamic, spatially localized nature of DENV transmission [9,10] implies that suppression of transmission in treated clusters could influence transmission in neighboring control clusters, thereby reducing incidence in both trial arms. Hereafter, we refer to each of these three forms of bias as “human movement,” “mosquito movement,” and “transmission coupling,” respectively.

In their per-protocol analyses, Utarini *et al*. [7] acknowledged the potential effects of human and mosquito movement in their per-protocol analysis, and by incorporating recent travel and *Wolbachia* prevalence into their efficacy calculations did not detect a difference in efficacy from that estimated in the intention-to-treat analysis. Nevertheless, the analysis of the AWED trial by Utarini *et al*. [7] did not account for transmission coupling, and they noted that follow-up analyses were needed to further explore the potential for bias due to human and mosquito movement.

Understanding the magnitude of such biases is important when seeking to extrapolate the impact of interventions across contexts. Such extrapolation has been recently undetaken for the RTS,S/AS01 vaccine [11,12] and the endectocide ivermectin [13] for malaria. If failing to account for such transmission dynamics contributes to an underestimated biological effect of *Wolbachia* on DENV, we risk incorrectly assessing its broader impact. Given the myriad intervention options available to public health officials for dengue control [14], it is important for the potential impacts of each to be understood as well as possible.

In this study, we used a mathematical model of DENV transmission to gain insight into the possible magnitudes of the three aforementioned sources of bias. Our approach involved translating model inputs of the basic reproduction number (*R*_0_), the spatial scale of human movement (*b*), and the proportional reduction in *R*_0_ afforded by *Wolbachia*-infected mosquitoes (*ε*) into outputs of the infection attack rate (IAR) in control and treatment arms of a trial, in accordance with a seasonal, two-patch susceptible-infectious-recovered (SIR) model [15]. We used the outputs of IAR in treatment and control arms (*IAR*_*t*_ and *IAR*_*c*_, respectively) to obtain an estimate of the odds ratio (OR) of infection and, thereby, an estimate of the efficacy of the intervention, *Eff* = 1 - *OR*. We constructed six different model versions for estimating efficacy, each of which includes different combinations of the three biases, all of them, or none of them. Henceforth, we refer to the efficacy observed in the AWED trial as “observed efficacy,” and the efficacy estimated by a given model and *ε* as “estimated efficacy.” Finally, we quantify each bias as the difference in the efficacy estimated by a model including that bias and a model which does not include that bias (see *Methods* for more details of our methods).

## Results

We assumed a checkerboard pattern of control and treatment arms of 1 km^2^ to approximate the design used in the AWED trial, which covered the entire city of Yogyakarta, with neighboring areas assigned to one arm or another in an (approximately) alternating pattern (Fig. 1A) [7], and assume that individuals are evenly distributed within each cluster such that they have no internal spatial structure. The time that humans spend away from their home is assumed to follow a Laplace distribution (Fig. 1A, top right), which takes a single parameter, *b*, that we refer to as the scale of human movement. By assuming that individuals are evenly distributed within each cluster, we can then estimate the average proportion of time that individuals in each trial arm spend in their own arm (*ρ*_*tt*_ and *ρ*_*cc*_) and in the opposite arm (*ρ*_*tc*_ and *ρ*_*ct*_ – see the *Apportionment of time at risk* section in *Methods* for details). Larger values of *b* imply that people spend less time in their allocated arm, and for large values of *b* individuals spend roughly equal amounts of time in both arms (Fig. 1B).

**Fig. 1:**
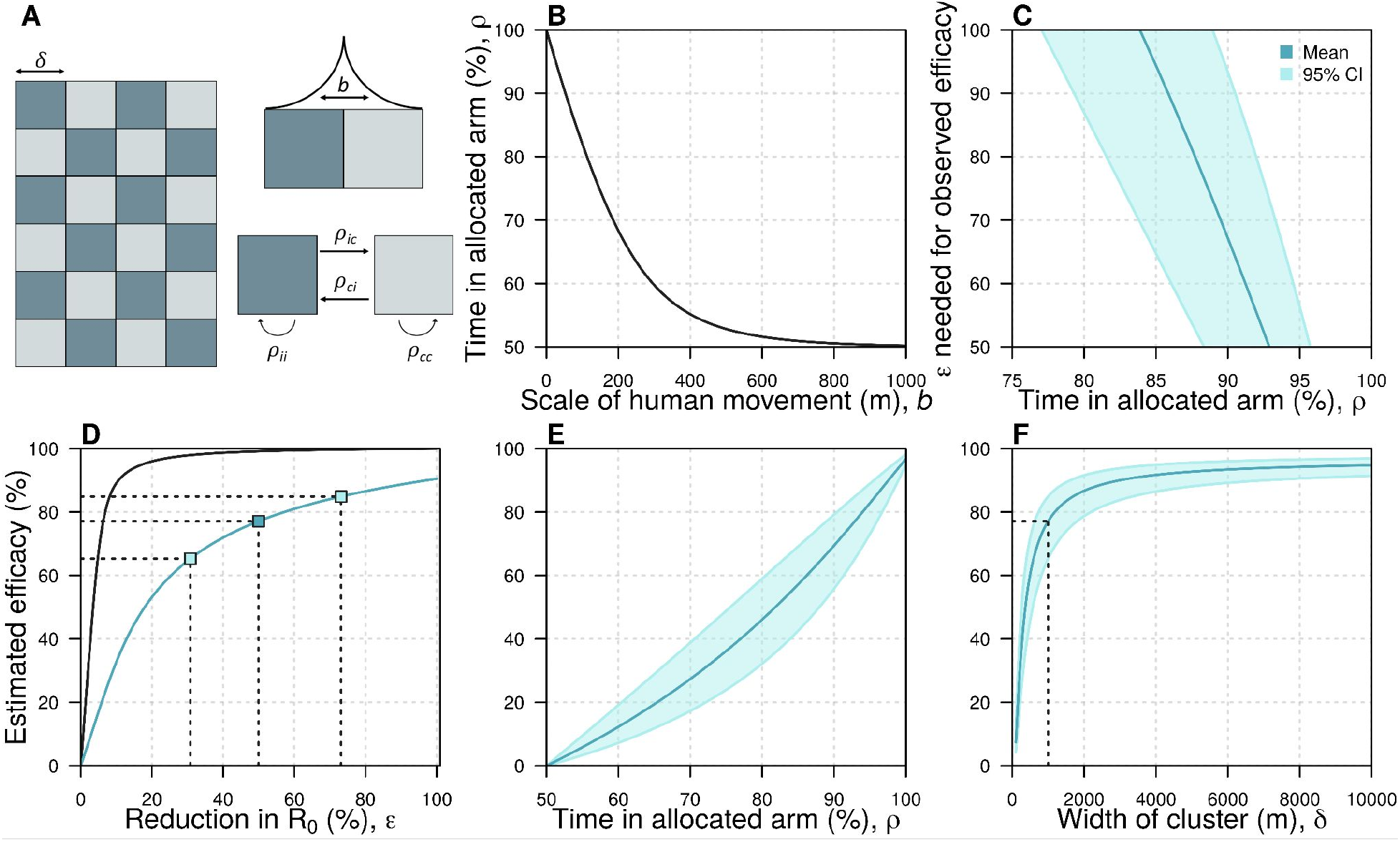
The spatial scales of transmission and trial design. A: Idealized trial design. We used a checkerboard pattern to approximate the design of the AWED trial of Wolbachia-infected mosquitoes to control dengue [7]. ρ_ij_ represents the amount of time an individual who lives in arm i spends in arm j, where i and j can represent either control (c) or treatment (i). b describes the scale of human movement. B: The relationship between the scale of human movement and the amount of time individuals spend in clusters of the same type as their home cluster. C: The relationship between the reduction in R_0_ (ε) required to reproduce the observed efficacy in the AWED trial and the time people spend in their allocated arm. In this panel and panels E and F, the dark blue line corresponds to the observed mean estimated in the AWED trial whereas the light blue line and shaded region correspond to the 95% confidence intervals. D: The relationship between ε and the estimated efficacy when b = 60 m. The black line shows the theoretical relationship between a reduction in R_0_ and observed efficacy, assuming no mosquito movement and no human movement between arms. The blue line shows this relationship if we include these two factors as well as the effect of transmission coupling. The dark and light blue squares indicate the mean and the 95% confidence interval respectively of the observed efficacy in the AWED trial and the corresponding reduction in R_0_. E: The relationship between the amount of time people spend in their allocated arm and the estimated efficacy. F: The relationship between the size of the clusters and the estimated efficacy. The dashed line indicates the estimated efficacy at the baseline cluster size (1000m). In all panels, parameters are at their baseline given in Table S1 unless otherwise stated.

The relationship between the efficacy estimated by the model with all three forms of bias (the estimated efficacy) and the reduction in *R*_0_ (*ε*) was dependent on the amount of time people spent in their allocated arm (Fig. 1C)—the less time individuals spent in their allocated arm, the higher the reduction in *R*_0_ that was needed to recreate the observed efficacy from the AWED trial. If individuals spent less than 83.9% of their time in their allocated arm, it was impossible to generate the observed efficacy (77.1%), as that would have implied that *ε* exceeded 1. Assuming that individuals spent 92.9% of their time in their allocated arm (i.e., *ρ*_*ii*_ *=* 92.9%, corresponding to *b* = 36.9 m – see the *Spatial Scale of Human Movement* section in *Methods* for details and justification), we found that the observed efficacy (77.1% [95% CI: 65.3% - 84.9%]) corresponded to an *ε* of 49.9% (95% CI: 30.8% - 73.1%) (Fig. 1D, blue line). If we instead assumed that there was no movement between trial arms, we observed that much smaller values of *ε* were needed to explain the observed efficacy (6.3% [95% CI: 4.8% - 8.1%]). The difference between these estimates provides an indication of the extent of bias introduced by assuming that humans and mosquitoes remain in their allocated arms, when they in fact do not (Fig. 1D).

When we fixed *ε* to the value that reproduces the observed efficacy in the AWED trial and increased human movement between arms by increasing *b*, the estimated efficacy by the model accounting for all three forms of bias decreased (Fig. 1E). For example, increasing the average distance in one direction between transmission pairs (*b*) from 36.9 m to 70 m caused a relative reduction of 20.0% in estimated efficacy, highlighting the sensitivity of efficacy to the spatial scale of human movement. This effect occurs for two reasons: first, as people spend less time in their allocated arm, the proportion of time that people spend under the intervention becomes more similar between arms; and secondly, in the presence of transmission coupling, a reduction in prevalence in the intervention arm reduces transmission in the control arm more as people spend less time in their allocated arm. Relatedly, estimated efficacy depended on the dimensions of the trial clusters, which we set to 1 km^2^ by default (Fig. 1F). When we reduced the cluster dimensions to 500 m × 500 m, estimated efficacy dropped from 77.1% to 60.3%, representing a 21.8% relative reduction. This effect occurs because, as the cluster dimensions are reduced, people spend less time in their home cluster. Hence, the time spent in each trial arm approaches parity (i.e., 50%). Increasing cluster dimensions above 1 km^2^ had somewhat less of an effect on estimated efficacy. For example, increasing the cluster dimensions to 2 km × 2 km resulted in an estimated efficacy of 86.7%, a relative increase of 12.4%.

Our approach enabled us to directly and separately model each of the three potential sources of bias: (1) mosquito movement, (2) human movement, and (3) transmission coupling. Movement of *Wolbachia*-infected mosquitoes is modeled by including a time-varying level of coverage, and we assume that mosquito movement does not contribute to DENV transmission (See *Methods - wMel coverage*). When we assumed that *ε* was equal to 49.9%, allowing for mosquito movement but not human movement produced an estimated efficacy of 99.1%, because there was almost no transmission in the intervention arm in that case (Fig. 2A, Fig. S7). If we allowed for both mosquito movement and human movement, we observed a lower estimated efficacy of 93.6%. Although there was little transmission in the intervention arm in this case, individuals residing in the intervention arm could be infected in the control arm. Additionally, those assigned to the control arm experienced lower overall risk due to their time spent in the intervention arm. When we accounted for transmission coupling between trial arms alongside human and mosquito movement, thereby allowing for more transmission in the intervention arm, risk was the most similar across the trial arms of all scenarios, leading to the lowest estimated efficacy of 77.1% for an *ε* equal to 49.9%.

**Fig. 2:**
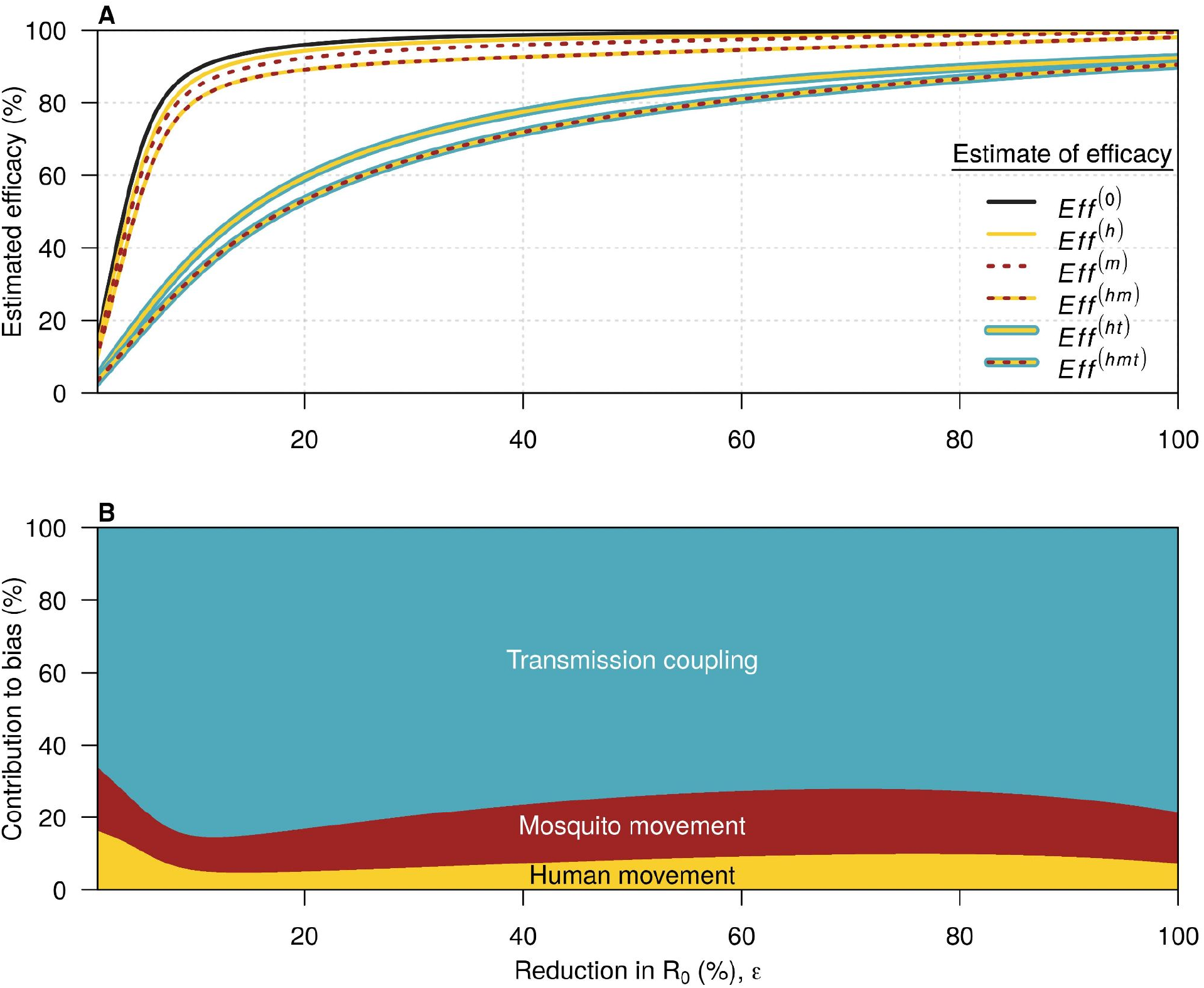
Sources of bias in efficacy estimates. In both panels, yellow refers to mosquito movement, red to human movement, and blue to transmission coupling. A: The relationship between the reduction in R_0_ (ε) and the estimated efficacy for the six possible models. The black line here is the relationship for a model with no human movement or mosquito movement. Where a line has more than one color, it represents the model which includes each of the types of bias represented by those colors. The difference between this line and each of the colored lines represents the bias introduced by not accounting for the features present in the model described by that colored line. B: the contribution of each source of bias to the total bias. Eff^(0)^ refers to the estimated efficacy from a model with none of the biases, Eff^(h)^ to the estimated efficacy from a model with human movement only, Eff^(m)^ to the estimated efficacy from a model with mosquito movement only, Eff^(hm)^ to the estimated efficacy from a model with human and mosquito movement, Eff^(ht)^ to the estimated efficacy from a model with human movement and transmission coupling, and Eff^(hmt)^ to the estimated efficacy from a model with all three biases.

We quantified total bias as *Eff*^*(hmt)*^ *- Eff*^*(0)*^, where *Eff*^*(hmt)*^ is the estimated efficacy under the model with all sources of bias and *Eff*^(0)^ is the estimated efficacy under the model without human or mosquito movement. We then computed the difference in the bias produced by pairs of models to decompose overall bias into each of its three sources (Fig. 2B, Fig. S8-9, see *Methods* for details). At the baseline *ε* of 49.9%, 17.6% of the total bias was attributable to mosquito movement, 8.3% to human movement, and 74.1% to transmission coupling. At all values of *ε*, the greatest source of bias was transmission coupling between trial arms. When *ε* was below a value of around 10%, the effective reproduction number at the start of the trial exceeded 1 in both arms. This value of *ε* varied slightly based on the model used (Fig. S8-9). If *ε* was below this critical value, increasing it in the context of coupled transmission reduced incidence in the control arm and caused smaller reductions in incidence in the intervention arm than if transmission had been uncoupled (Fig. S7, e.g. panels D vs. F). This implies that the bias introduced by transmission coupling increases as *ε* increases up to ∼10% under our model’s parameterization (Fig. 2B). Increasing *ε* past this point only leads to small reductions in incidence in the intervention arm in an uncoupled model, as incidence is already very low.

## Discussion

Our results highlight three sources of bias (human movement, mosquito movement, and transmission coupling) that arise in large, cluster-randomized, controlled trials of interventions against mosquito-borne diseases, and have implications for how to mitigate these biases. Biases arising due to human movement and mosquito movement are typically able to be addressed through careful statistical analysis of trial data or in the design of the trial [8]. For instance, in the per-protocol analysis of the AWED trial, Utarini *et al*. accounted for these two forms of bias by combining self-reported recent travel and local *Wolbachia* prevalence into an individual-level *Wolbachia* exposure index [7]. Comparing groups with the highest and lowest *Wolbachia* exposure did not lead to higher efficacy estimates than their primary analysis. Another approach to addressing contamination involves describing the effectiveness of the intervention at the boundary between clusters using a sigmoid function [16–18]. Our results suggest that failure to take steps such as this to account for human and mosquito movement would typically lead to underestimated efficacy, while failure to account for transmission coupling would lead to an even greater underestimate, particularly at intermediate reductions in *R*_*0*_.

Bias arising from human and mosquito movement could also be mitigated at the stage of planning the trial. The classical design to achieve this is the ‘fried-egg’ design, in which a treated buffer-zone is placed between intervention and control clusters [19]. A more recently proposed approach involves excluding a subset of clusters from the trial completely, thereby increasing the distance between clusters and leading to disconnected clusters at less risk of contamination [20]. While both of these approaches do mitigate the risk of contamination directly, they also necessitate a larger trial area and may be logistically infeasible in a trial taking place in a single city, as was the case for the AWED trial. Another approach could include reducing the number of clusters, but keeping the total area fixed, leading individuals to spend more time in their assigned arm and reducing mosquito movement by reducing the boundary between clusters. Our results show that the efficacy estimated from cluster-randomized, controlled trials of interventions against mosquito-borne diseases is highly sensitive to cluster size (Fig. 1F). Had the dimensions of the clusters in the AWED trial been much smaller, then the estimated efficacy may have been substantially lower. However, having fewer, yet larger clusters would likely introduce new biases by making the arms less comparable, which may not be an acceptable trade-off.

While bias due to human and mosquito movement can be mitigated through trial design and statistical methods, our results highlight a third source of bias, transmission coupling, that requires additional tools to fully address. Accounting for this bias first requires data on the spatial distribution of the intervention and on human movement, similar to that used in the supplementary analysis of the AWED trial. However, it also requires interfacing these data with a dynamical transmission model to account for the fact that, in the presence of movement between arms, incidence in each arm depends on prevalence in both arms [21]. Many common trial designs will lead to reduced bias due to transmission coupling – for instance by allocating a greater proportion of the trial area to the control arm, with small intervention clusters situated among larger control clusters so that transmission suppression in the intervention arm has less of a population-level effect. The ratio of area allotted to treatment and control would depend on many factors, including the expected strength of the intervention, the local force of infection, and logistical constraints such as the size and length of the trial. Utilizing a dynamical model synthesizing these factors in the design of a trial could aid in understanding how different designs might affect bias due to transmission coupling [21]. More work is needed to understand what types of spatial clustering patterns, among other features of trial design, would minimize this form of bias.

Although our modeling approach allowed us to account for different potential sources of bias and to attribute the total bias to each of those sources, it has at least four limitations. First, our model was deterministic, yet stochasticity could be important for a highly efficacious intervention with potential to reduce transmission to very low levels [22]. This simplification implies that our estimates are likely conservative, as these effects could increase the bias due to transmission coupling if a highly effective intervention increases the probability of transmission fadeouts. Second, our simple model does not reflect all of the complexities of DENV transmission. For example, we did not account for spatial heterogeneities in transmission or interactions between serotypes. Accurately quantifying the contribution of these effects to bias would require a more detailed model, but the qualitative results would likely be similar. Third, we did not calibrate our model to trial data, so incidence in our model may not reflect the actual incidence during the trial. However, our aim here was not to precisely quantify bias in the AWED trial, but rather to highlight some potential sources of bias in trials of that nature and to understand how these biases are influenced by transmission dynamics and human mobility. Moreover, our model was calibrated to actual incidence from past years in Yogyakarta, and so still reflects transmission typical of that location. It is also worth noting that an earlier version of the manuscript, which used a simpler static model based on epidemic attack rate formulae, had qualitatively similar findings [23]. Finally, we don’t account for heterogeneity between clusters, such as regions of the city with systematically higher mosquito abundance, or greater human movement, or within clusters, such as that transmission may be higher at the edge of control clusters.

In conclusion, without accounting for human movement, mosquito movement, and transmission coupling, the efficacy of *Wolbachia*-infected mosquitoes as an intervention to control dengue is likely to be underestimated. As the estimate of efficacy in the AWED trial was already very high (77.1% [95% CI: 65.3% - 84.9%]) [7] and, as we show, likely underestimated, *Wolbachia*-infected mosquitoes have potential to be a game-changing tool in the fight against dengue. Even as vaccines against dengue become available, a variety of vector control approaches are likely to remain key tools in the fight against dengue [2,14]. Although we focused our study on a trial of *Wolbachia*-infected mosquitoes, our findings are applicable to any efficacy trial of an intervention that has the potential to contaminate the control arm, such as gene drive mosquitoes or ivermectin as interventions against malaria [24,25]. As trials of these interventions continue, it will be important to learn what lessons we can from transmission dynamic modeling when designing and interpreting future trials to ensure that we understand the true promise of these interventions.

## Methods

### Transmission model

We simulated DENV transmission using a four-serotype, two-patch seasonal SIR model. In this model, fully susceptible individuals may become infected with any of the four serotypes. Once infected, individuals have an exponentially-distributed period of cross-immunity to all other serotypes with a mean of two years. Individuals with prior exposure to one or more serotypes but that are not currently in their period of cross-immunity are immune to the serotypes they have previously been infected with. We implicitly assume that all four serotypes circulate in equal proportions. Births and deaths are modeled so that the population size remains constant, and the mortality rate is the reciprocal of the mean life expectancy, taken from the United Nations World Population Prospects database [26]. The transmission parameter, *β*(*t*), varies seasonally according to a sine curve with a period equal to one year. The model equations are as follows, with parameter definitions and values given in Tables S1 and S2, and the model diagram is shown in Fig. S1.

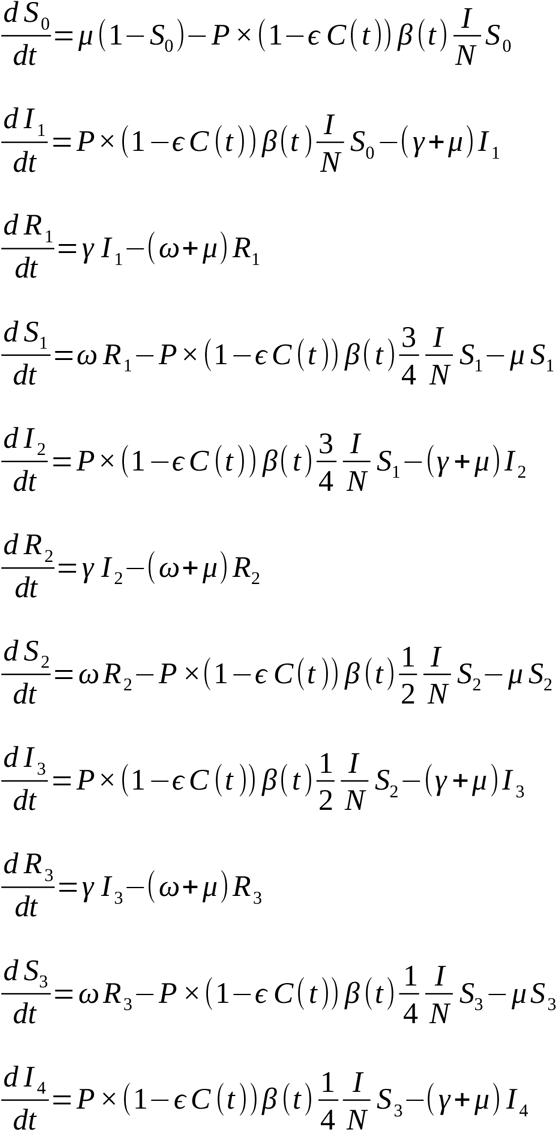

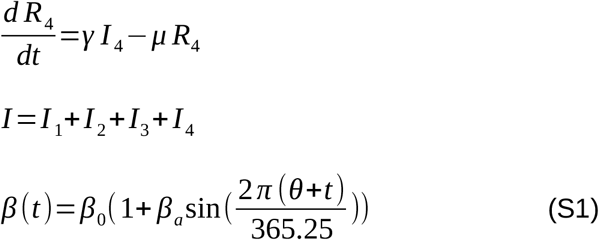

### Transmission model calibration

We calibrated the model to data on reported cases of dengue fever over a ten year period [29] (Fig. S2). We first averaged the monthly number of reported cases, to capture the average dynamics across the period. We ran the model for 100 years to reduce the influence of initial conditions, and then fitted model years 101-110 to the 10 average years from the data using maximum likelihood. We used a Poisson likelihood function,

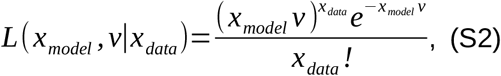

where *x*_*model*_ is the number of infections per month predicted by the model and *x*_*data*_is the number of cases per month in the data.

### Efficacy models

Let *ε* represent the effectiveness of the intervention, defined as the proportional reduction in the pre-intervention basic reproduction number, *R*_0_, when the intervention is applied at full coverage in a treatment cluster. Hence, in the absence of human or mosquito movement,

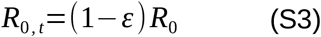

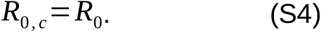

Our interest is in quantifying the infection attack rate (IAR), *π*, within each cluster during a trial. To do this, we simulate the model for two years, and calculate the infection attack rate in each arm during that time. We estimate the initial proportion in each compartment by first simulating the model for 100 years. We do this with six different models that include combinations of three different types of bias: human movement between arms, mosquito movement between arms, and transmission coupling between arms. The six resulting models are described below (note that transmission coupling can only occur in the presence of human movement). Each model is defined by different values for P and *C*(*t*).

#### 1. No bias

In the absence of contamination from human movement or mosquito movement between arms, we can essentially use equations (S3) and (S4) to describe the reproduction in each arm. This amounts to setting *C*(*t*) = (1, 0) and P = I, the identity matrix.

#### 2. Bias from mosquito movement

We represent the coverage of the intervention—i.e., the proportion of *Wolbachia*-infected mosquitoes—in the two arms with *C*_t_(*t*) and *C*_c_(*t*). In the case of mosquito movement, there may be non-zero coverage of intervention in the control arm (i.e., *C*_c_>0), and less than 100% coverage in the treatment arm (i.e., *C*_*t*_ < 1). Hence we set *C*(*t*) = (*C*_*c*_(*t*), *C*_*t*_(*t*)) and P = I.

Here we are assuming that movement of mosquitoes between trial arms does not directly contribute to DENV transmission via movement of DENV-infected mosquitoes. This discrepancy can be reconciled by the fact that the spread of dengue virus occurs within a single mosquito generation, whereas the spread of *Wolbachia* occurs over the course of multiple generations.

#### 3. Bias from human movement

Let ϱ_ij_ represent the ij^th^ element of P,—i.e., the proportion of the total time at risk that a resident of cluster *i* spends in cluster *j*. To account for human movement, but no transmission coupling, we set *C*(*t*) = (ϱ_ct_, ϱ_tt_) and P = I. This is because in this scenario, the wMel coverage in the treatment arm is 1, and in the control arm is 0, so the experienced wMel exposure reduces to the time spent in the treatment arm.

#### 4. Bias from human movement and mosquito movement

We now have both human and mosquito movement, so we set

*C*(*t*) = (ϱ_cc_*C*_*c*_(*t*) + ϱ_ct_*C*_*t*_(*t*), ϱ_tc_*C*_*c*_(*t*) + ϱ_tt_*C*_*t*_(*t*)), and P = I. Note that, by definition, *ϱ*_tt_ + *ϱ*_tc_ = 1 and *ϱ*_cc_ + *ϱ*_ct_ = 1.

#### 5. Bias from human movement and transmission coupling

Thus far, we have assumed that transmission in each arm is only a function of prevalence in that arm, and not in the other. To relax this assumption, we couple transmission between the two arms by varying P. In the presence of human movement but not mosquito movement, we set C = (0, 1) and P = (ϱ_cc_, ϱ_ct_; ϱ_tc_, ϱ_tt_).

#### 6. Bias from human movement, mosquito movement, and transmission coupling

Finally, we include all three forms of bias by again setting P = (ϱ_cc_, ϱ_ct_; ϱ_tc_, ϱ_tt_), and C = (*C*_*c*_(*t*), *C*_*t*_(*t*)).

### Efficacy calculation

The ratio of the IARs in the treatment and control clusters is an infection risk ratio. However, the AWED trial based their efficacy calculations upon an odds ratio [7], with symptomatic, virologically-confirmed dengue as the end point. That is, efficacy in the trial was computed as 1-*p*_t_*n*_c_/*p*_c_*n*_t_, where *p*_i_ and *n*_i_ represent enrolled test-positives and test-negatives, respectively, in trial arm *i*. To generate a comparable quantity, we computed the efficacy according to model *x* as

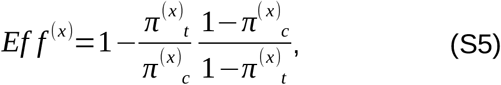

where *π* ^(x)^_*i*_ is the infection attack rate in trial arm *i*∈{*c,t*} for model *x*∈{*0,h,m,hm,ht,hmt*}. Here, we are assuming that the ratio of infections to enrolled test-positives does not differ between arms (i.e., *p*_i_=*k*_p_*π* _i_ for *i*∈{*c,t*}) and similarly the ratio of those uninfected to enrolled test-negatives also does not differ between arms (i.e., *n*_i_=*k*_n_(1-*π*_i_) for *i*∈{*c,t*}). If either of these assumptions were violated, for instance if the intervention affected either the proportion of dengue infections that were symptomatic, then our estimate of efficacy would be less comparable to the estimate used in the AWED trial.

### Bias calculation

We calculated the bias due to a particular source as the difference in the efficacy between a model with that source of bias and a model without that source of bias. As biases appear in multiple models, this led to three ways to embed the models, and three corresponding ways to quantify each bias. The three embeddings are: A) no bias → mosquito movement → human movement + mosquito movement → full model; B) no bias → human movement → human movement + mosquito movement → full model; and C) no bias → human movement → human movement + transmission coupling → full model. The difference between efficacy estimates for adjacent models in an embedding will lead to an expression for the bias which differs between the two models. Hence, the three possible ways to calculate each of the three sources of bias yields

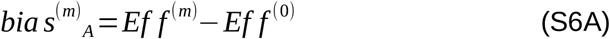

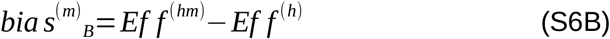

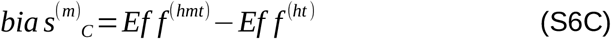

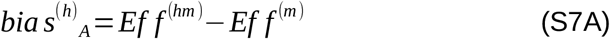

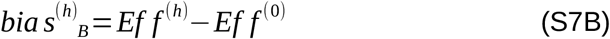

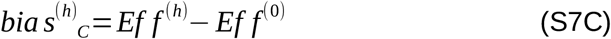

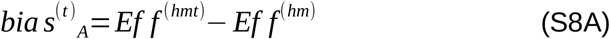

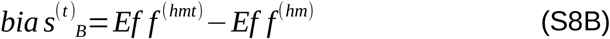

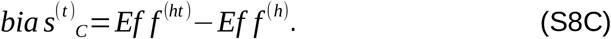

We then calculate the average total bias caused by each source of bias as

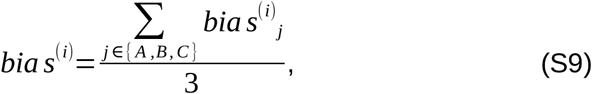

where *i*∈{*h,m,t*}. Note that *bias*^*(t)*^_*A*_=*bias*^*(t)*^_*B*_ and *bias*^*(h)*^_*B*_=*bias*^*(h)*^_*C*_, but it is necessary to include each as a separate term so that each of the three model embeddings is included equally.

### Model Parameterization

#### Apportionment of Time at Risk

We considered a checkerboard arrangement for the treatment and control clusters in a trial across a two-dimensional landscape (Fig. 1A, Fig. S10). Under this scenario, we assume that the population density per unit area is constant and that transmission potential, as captured by *R*_0_, is homogeneous across the landscape prior to initiation of the trial.

At the core of this derivation is the assumption that the location where an individual *j* resides who was infected by an individual *i* is determined by an isotropic transmission kernel, *k* (|*x*_*i*_−*x* _*j*_|,|*y*_*i*_− *y* _*j*_|), where *x* and *y* are the spatial coordinates for the residence of each of *i* and *j*. We use a Laplace distribution with marginal density functions for each of the *x* and *y* coordinates,

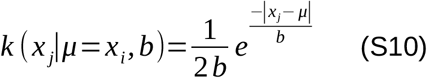

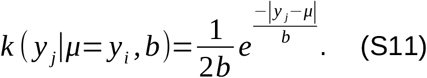

where *µ* is the location parameter and *b* is the scale parameter [30]. The scale parameter *b* is equal to the average distance in one direction between the locations where infector and infectee reside.

Under the checkerboard arrangement, we considered alternating squares of width *δ* corresponding to treatment and control clusters within a contiguous urban area (Fig. 1A). Although any such area would have borders in reality, we ignored any possible edges effects and assumed that the extent of interactions between squares of type *t* and *c* in the interior of the checkerboard provide a suitable characterization of overall interaction between individuals residing in *t* and *c*, as summarized by *ρ*_*tt*_ and *ρ*_*cc*_. Because the area and arrangement of *t* and *c* squares are identical, *ρ*_*tt*_ = *ρ*_*cc*_ and *ρ*_*tc*_ = *ρ*_*ct*_ (Fig. 1A).

We approach this problem by first calculating the proportion of time at risk that an individual *i* residing on a line within in an interval of width *δ*=*µ*_r_-*µ*_l_ experiences in an adjacent interval of width Δ. Let the former interval span [*µ*_l_,*µ*_r_] and the latter interval span [*µ*_r_,*µ*_r_+Δ]. If *i* resides specifically at *µ*, then the proportion of its time at risk in the other interval is

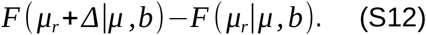

where *F*(·) is the Laplace distribution function. To average across all individuals *i*, we can integrate according to

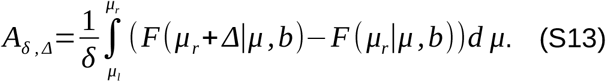

which gives the proportion of time in the interval of length Δ for an individual who resides in the interval of length *δ*. Given that the Laplace distribution function is *F*(*x*|*µ,b*)*=*1-½exp(-(*x*-*µ*)/*b*) when *x* >*µ*, eqn. (S25) evaluates to

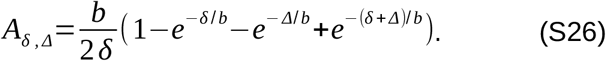

We can quantify the proportion of time at risk in the interval of width *δ*for individuals who reside there as

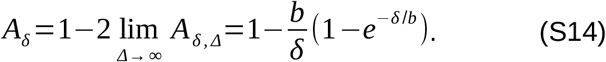

We also need to calculate the time at risk in a non-adjacent interval of width *δ*_3_ whose edge is spaced distance *δ*_2_ away from the nearest edge of the interval where the individual resides, which has width *δ*_1_. Applying similar reasoning as in eq. (S25), we obtain

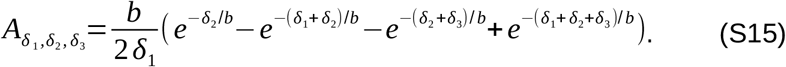

We can calculate the proportion of time spent in like squares by applying the probabilities used to calculate the proportions of time at risk for residents who live under treatment or not. Going out three layers from a focal square (Fig. S10), the proportion of time spent in like squares is

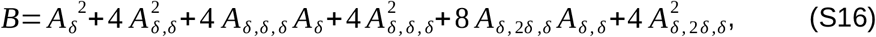

and the proportion of time spent in unlike squares is

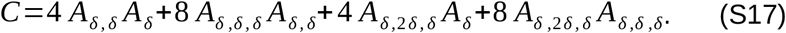

The total proportion of time under treatment or not is then

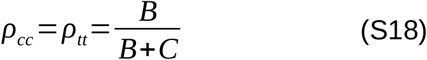

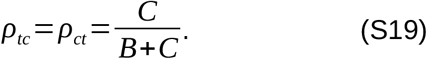

Hence, for a checkerboard arrangement of clusters, the proportion of time which each individual spends in each arm of the trial is uniquely determined by the width of each cluster (*δ*) and the scale of human movement (*b*).

#### Calculation of Initial Susceptibility, force of infection, and R_0_

To obtain an estimate of initial susceptibility, we followed ten Bosch *et al*. [31] and calculated the proportion of the population exposed to *n* serotypes, ∀*n*∈{0,1,2,3,4}, as a function of age. Following ten Bosch *et al*. [31], we defined *e*_i_(*a*) as the proportion of individuals of age *a* that have been exposed to *i* serotypes and *r*_i_(*a*) as the proportion of individuals of age *a* experiencing temporary heterologous immunity following exposure to *i* serotypes. The dynamics of how individuals progress through these classes as they age follows

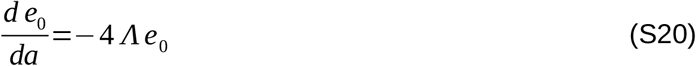

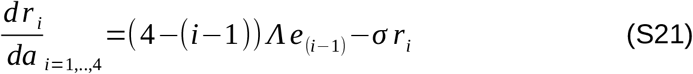

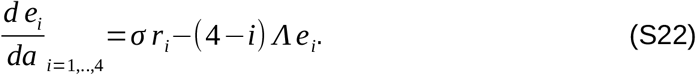

In eqs. (S33-S35), Λ = 0.0457 is the force of infection, and *σ* is the rate at which individuals lose heterologous immunity, which we set to 0.5/yr [31].

We computed the proportion of the population in Yogyakarta, Indonesia that is of age *a* using estimates from the United Nations World Population Prospects database [32] and computed the proportion of the population that is susceptible to their (i+1)th infection as

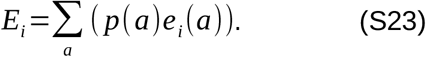

It follows that initial susceptibility is equal to

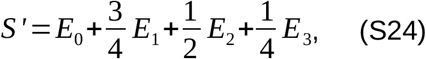

provided that the force of infection for each serotype has been constant over time. For the assumed values of Λ and σ, S’ = 0.341 for Yogyakarta, Indonesia.

We used data on seropositivity by age from Yogyakarta [29] to estimate the mean annual force of infection using the above catalytic model (Fig. S3). This led to an estimate of the mean annual per-serotype force of infection of 0.0457.

To estimate R_0_ from Λ and S’, we use the formula:

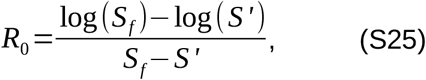

where *S*_*f*_ is the proportion susceptible after an outbreak. Here we estimate *R*_*0*_ based on one season’s transmission, i.e., *S*_*f*_ =*S’* exp(−4 *Λ*), yielding *R*_0_ = 3.21. We incorporate this estimate of *R*_*0*_ into the transmission model by assuming that the mean value of β(*t*) (i.e. β_0_) is related to *R*_*0*_ by *β*_0_=*R*_0_ *γ*, i.e. we assume that *R*_*0*_ represents the number of secondary infections in a fully susceptible population in the absence of seasonality. It is likely that this leads to an overestimate of β_0_, though our model still accurately recreates the typical epidemic peaks and troughs of Yogyakarta (Fig. S2).

#### Spatial Scale of Human Movement

Our calculations of the apportionment of time at risk depend upon a value of *b*, the spatial scale of human movement, a quantity that is challenging to parameterize. To do this, we first estimate the relative risk (RR) of 100% wMel coverage compared to 0% wMel coverage, based on the per-protocol analysis in Utarini *et al*. In that analysis, the authors estimated a weighted wMel exposure level based on human movement diaries and local wMel frequency over time. This is essentially the product of the *wMel* frequency in a location and the amount of time an individual spent there, and then summed over all of the locations at which that individual spent time. They then binned individuals into five equal width groups based on their exposure index and calculated the RR of infection compared to the lowest exposure group (Fig. S4). To estimate the RR of 100% exposure compared to 0%, we fit a logistic curve to the binned RR values (using the midpoints of each bin) and calculate the RR of 100% compared to 0%. This yields a RR of 0.18, or equivalently an efficacy of 82%.

To inform our selection of *b*, we compared the model with all of the biases included to the one with only human movement and transmission coupling. We then selected a value of *b* which enabled us to select a single value of ε that would lead to 77% efficacy in the full model, and 82% efficacy in the model with human movement and transmission coupling (Fig. S5). This yielded a value of *b*=36.9m, corresponding to *ρ*_*tt*_ =*ρ*_*cc*_=0.929 and *ρ*_*tc*_=*ρ*_*ct*_=0.071 for the checkerboard arrangement.

### wMel coverage

When modeling mosquito movement between trial arms, we use data on the time-varying wMel coverage in each trial arm from the AWED trial [7]. We average across clusters within each arm to find the average coverage over time. As we don’t explicitly model mosquitoes or their movement, these averaged time series are then used directly in the model. They are shown in Fig. S6 and are represented in the model by *C*(*t*) = (*C*_*c*_(*t*), *C*_*t*_(*t*)), where *C*_*c*_(*t*) is given in the left panel and *C*_*t*_(*t*) in the right. When mosquito movement is not modeled, *C*(*t*) = (0, 1), for all *t*.

## Data Availability

All model code is available at https://github.com/scavany/awed_trial_modeling

## Data availability

All code and other files to reproduce our results is available at: https://github.com/scavany/awed_trial_modeling/

## Supplementary Figures

**Table S1.**
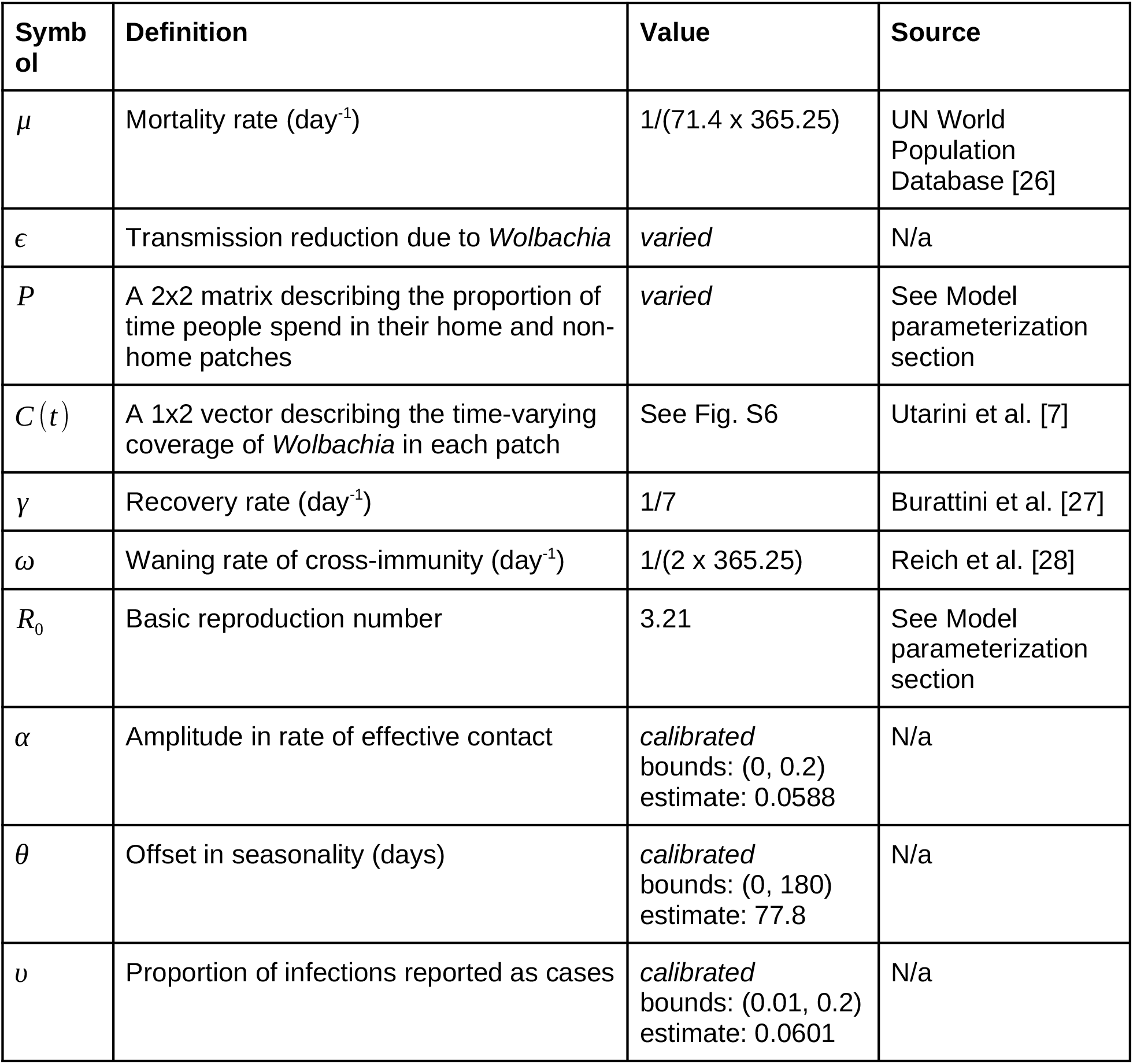
Model parameter values

**Table S2.**
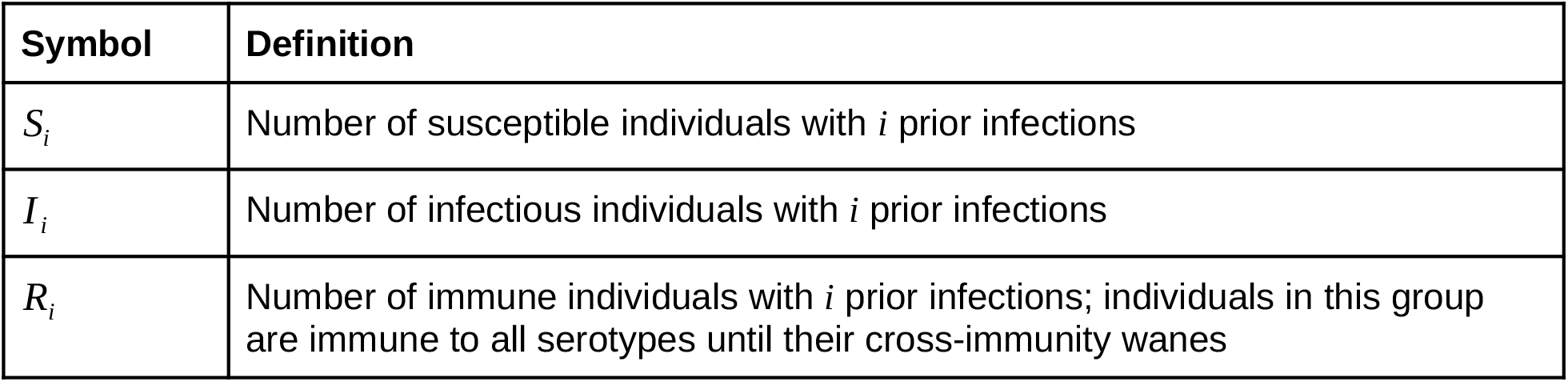
All state variables are 1×2 vectors describing the number in each of the two patches.

| Symbol | Definition |
| --- | --- |
| $S_i$ | Number of susceptible individuals with $i$ prior infections |
| $I_i$ | Number of infectious individuals with $i$ prior infections |
| $R_i$ | Number of immune individuals with $i$ prior infections; individuals in this group are immune to all serotypes until their cross-immunity wanes |

**Fig. S1:**
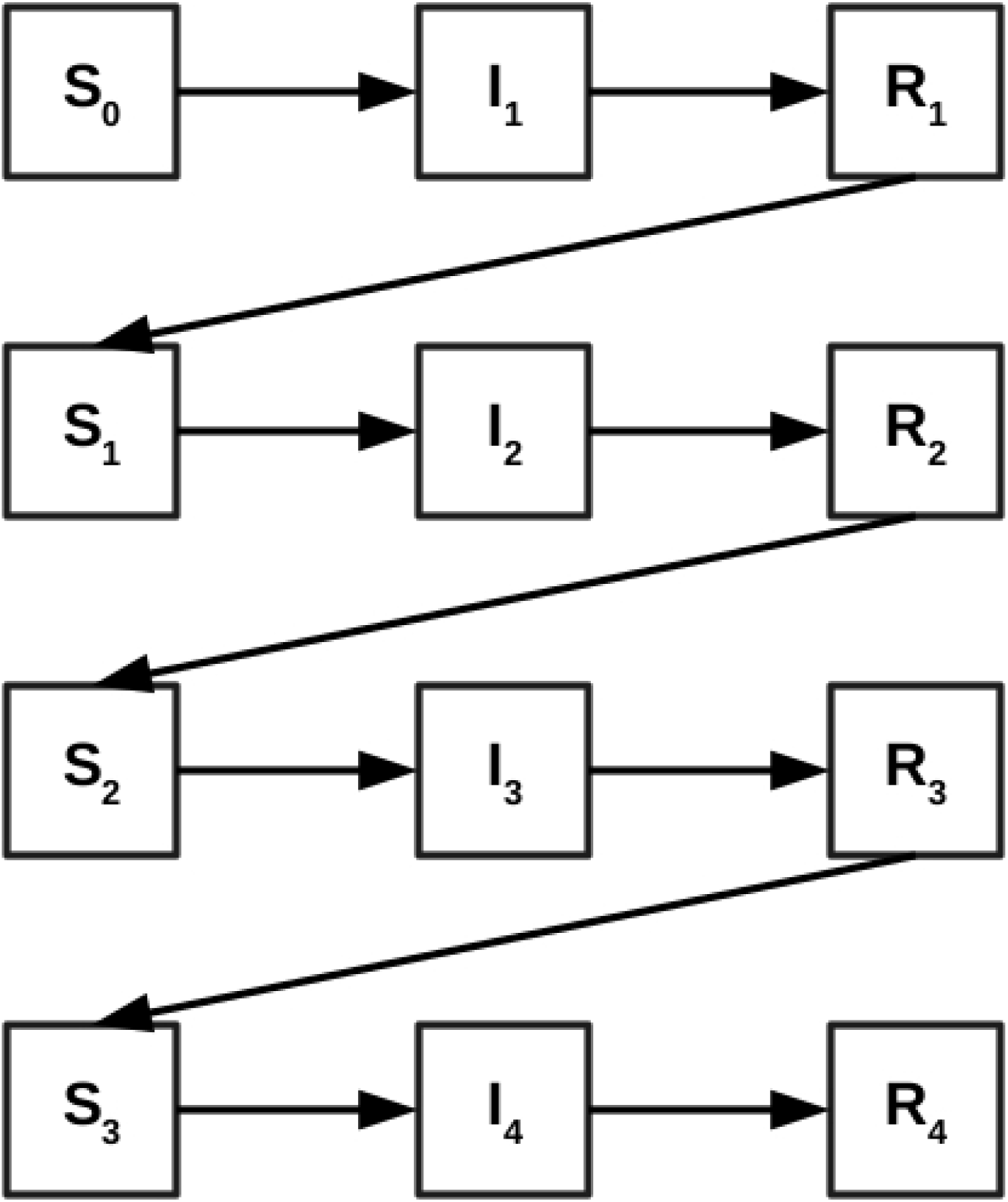
Model diagram. The superscripts refer to the number of times individuals in that compartment have been infected. Susceptible individuals (S_i_) experience a reduced force of infection according to the number of prior infections they have experienced. We assume all serotypes circulate equally. Following infection, individuals experience a temporary period of immunity to all serotypes (R_i_). Mortality occurs at an equal rate from all compartments and is not shown.

**Fig. S2:**
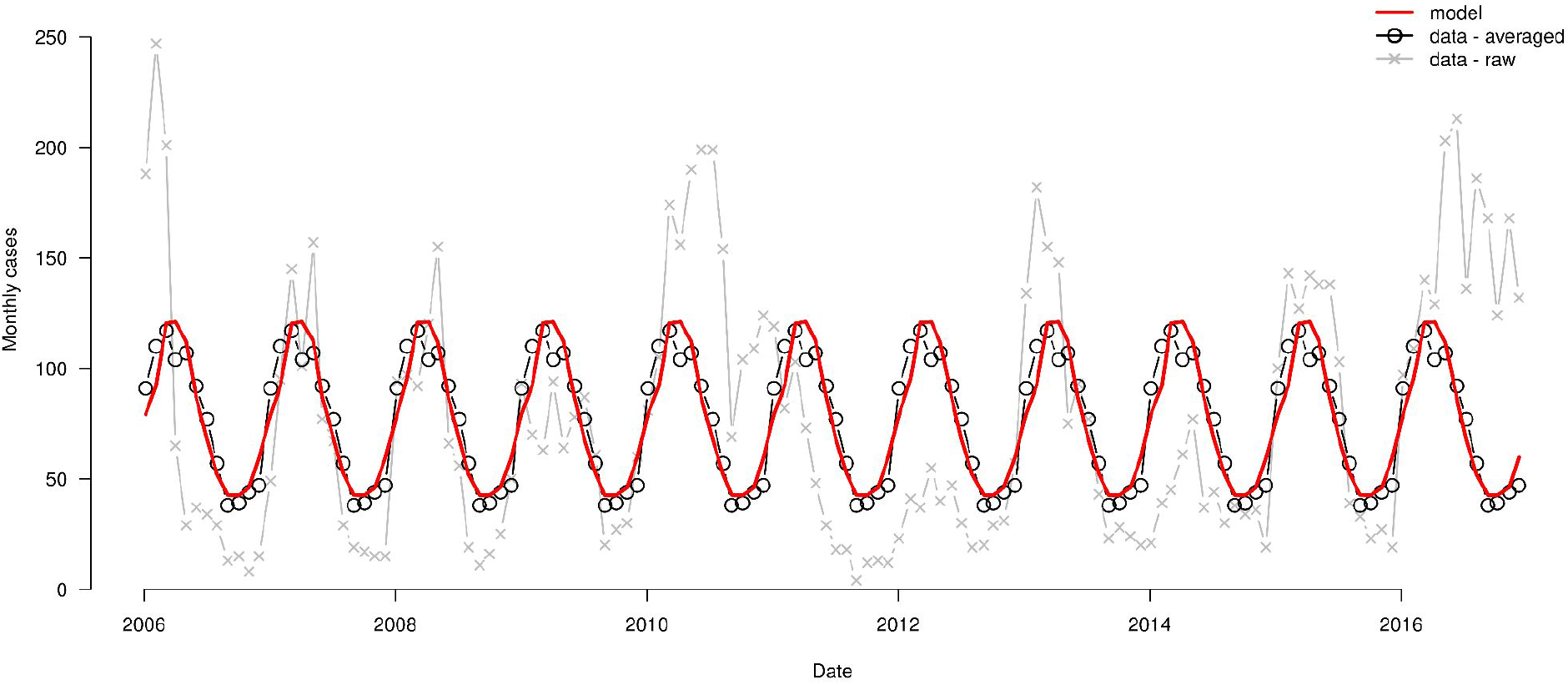
Model calibration. Calibration of seasonal SIR model to data on dengue cases from Yogyakarta. The faint red line and points show the data on the monthly number of cases from 2006 to 2017 in Yogyakarta, taken from Indriani et al. [29]. The solid red lines and points show this data average by month. The gray polygon shows the model calibrated to the average number of monthly cases.

**Fig. S3:**
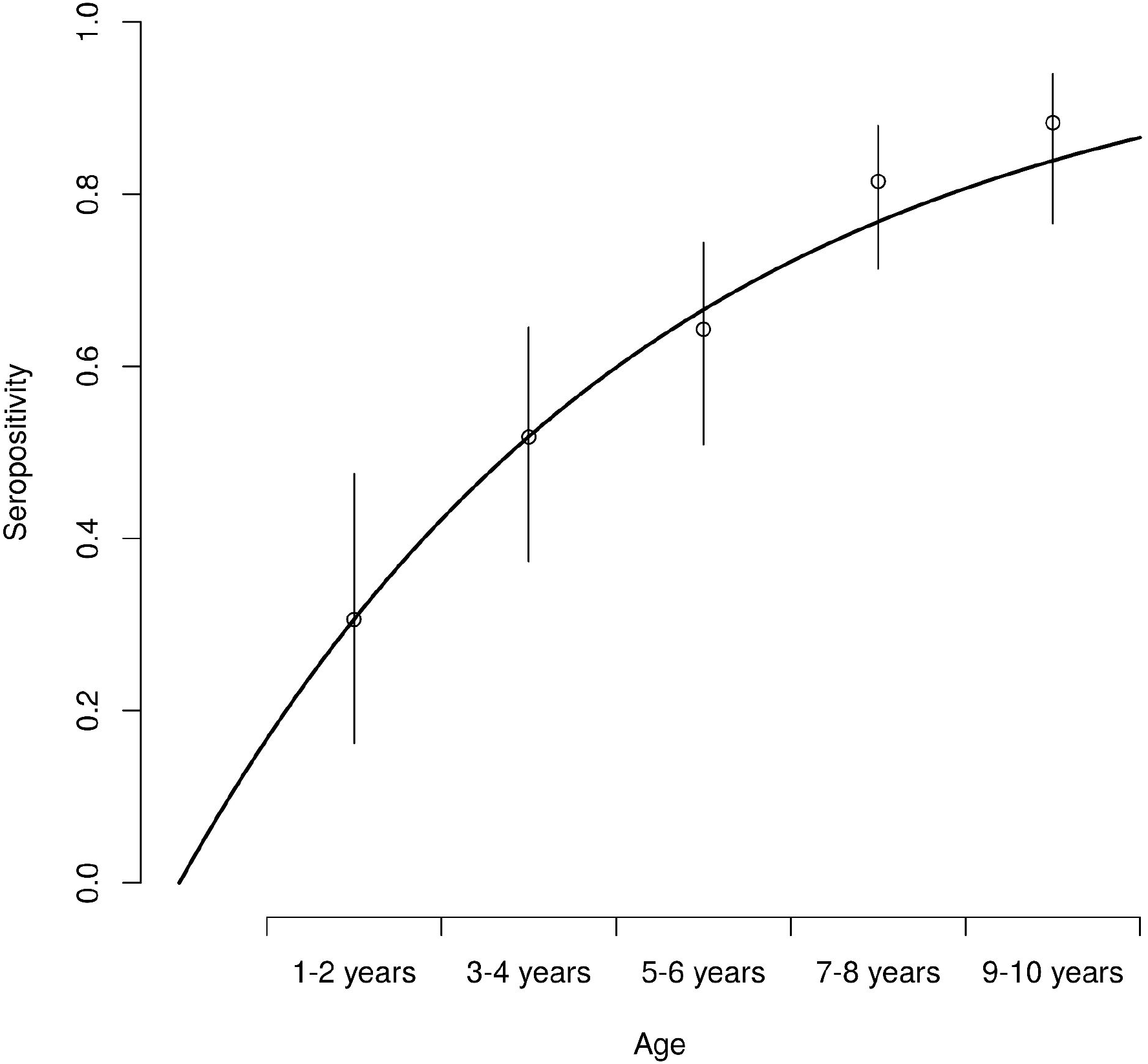
Force of infection estimation. The circles show the proportion of individuals that are seropositive by age group in Yogyakarta, and the thin vertical lines show the 95% binomial confidence intervals, both from Indriani et al. [29]. The thick black line shows the proportion that would be expected to be seropositive according to the catalytic model with a per-serotype force of infection of 0.0457.

**Fig. S4.**
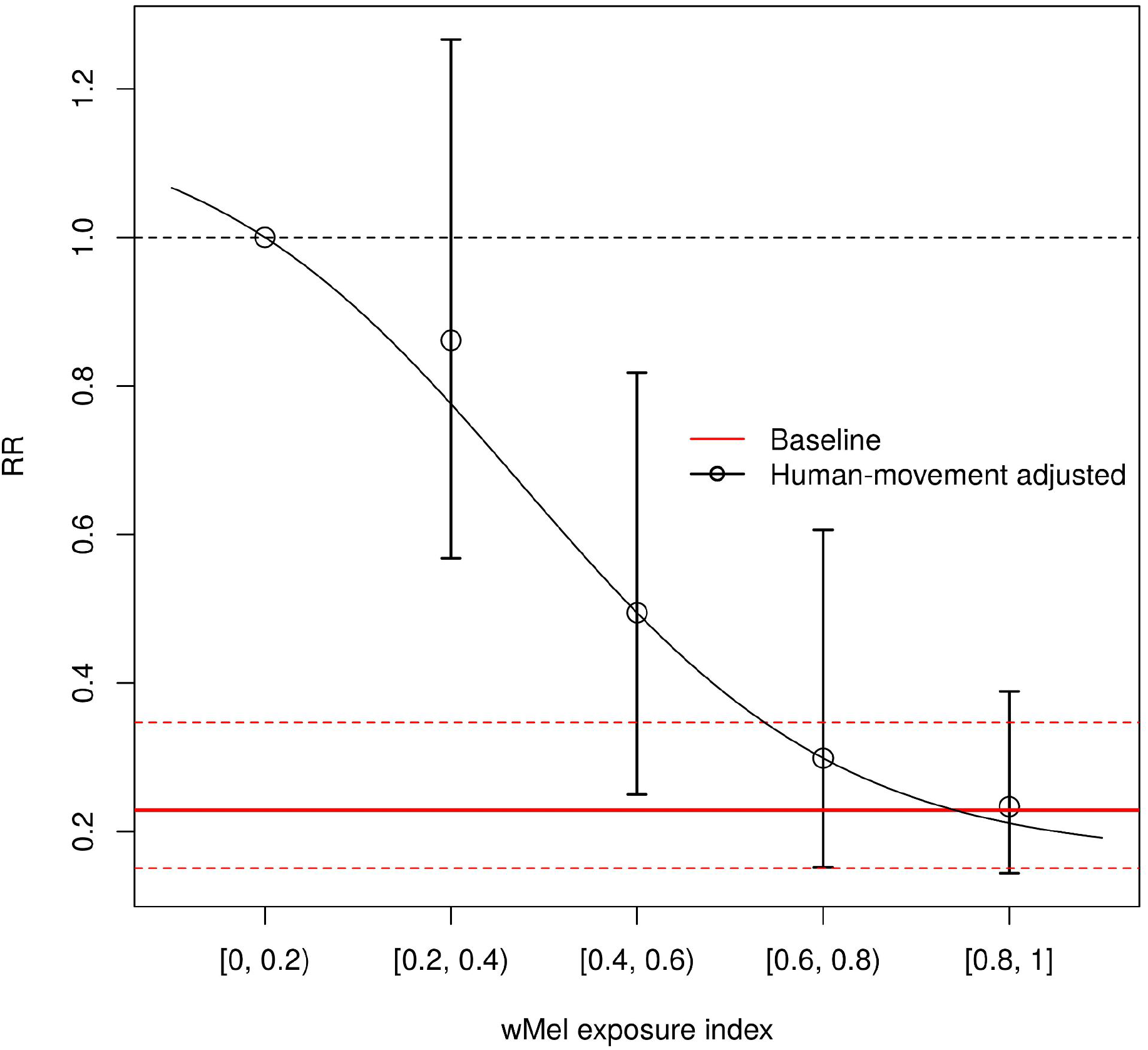
Efficacy adjusted for human movement. Circles and associated confidence intervals show the relative risk at different levels of the wMel exposure index compared to the [0, 0.2) group, according to the per-protocol analysis in Utarini et al. [7]. The horizontal red lines show the relative risk from the intention-to-treat analysis in the same paper. The dashed horizontal indicates a relative risk of 1. The black line indicates a logistic curve fit to the estimates of relative risk from the per-protocol analysis.

**Fig. S5:**
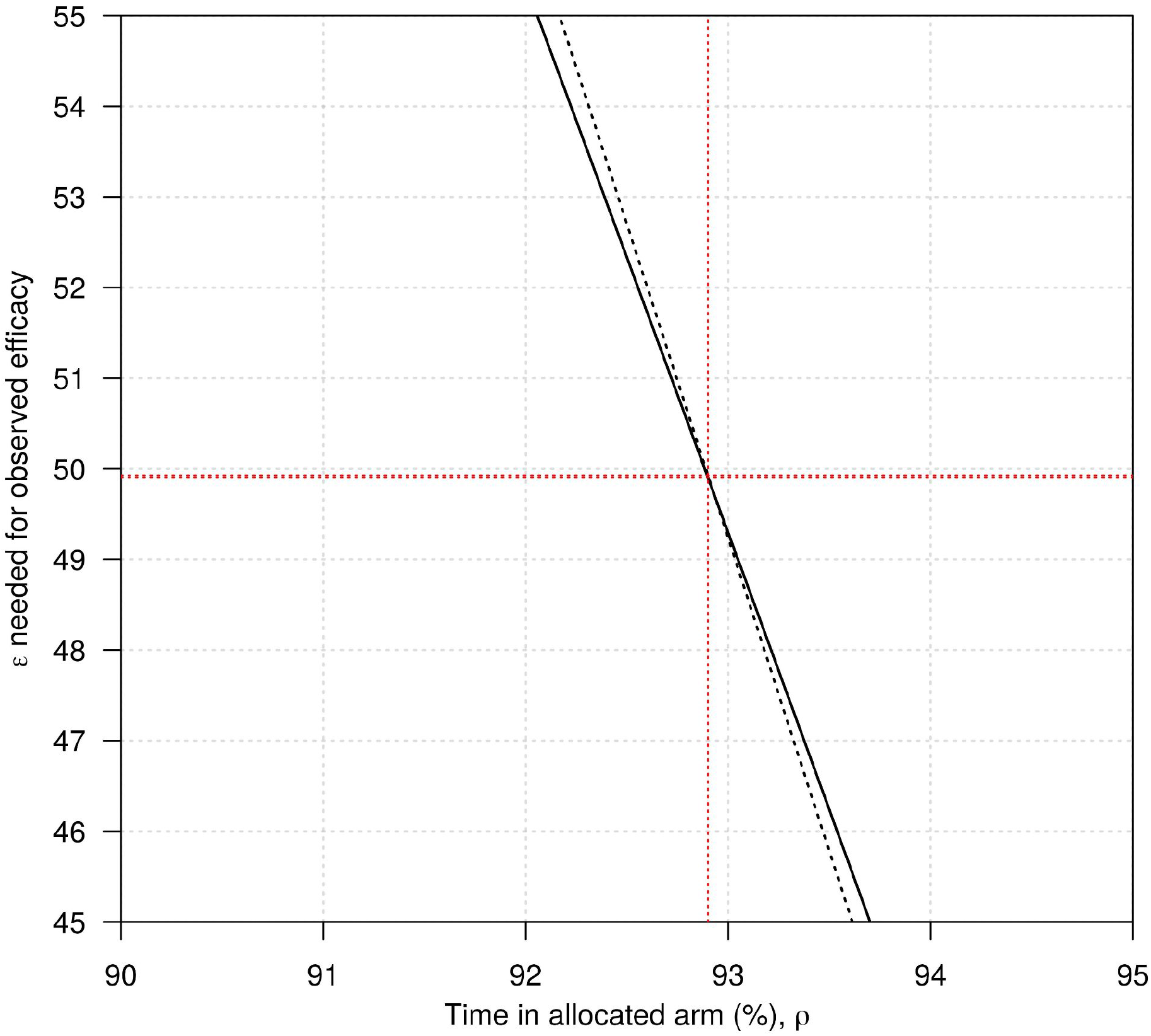
Estimation of scale of human movement.

**Fig. S6.**
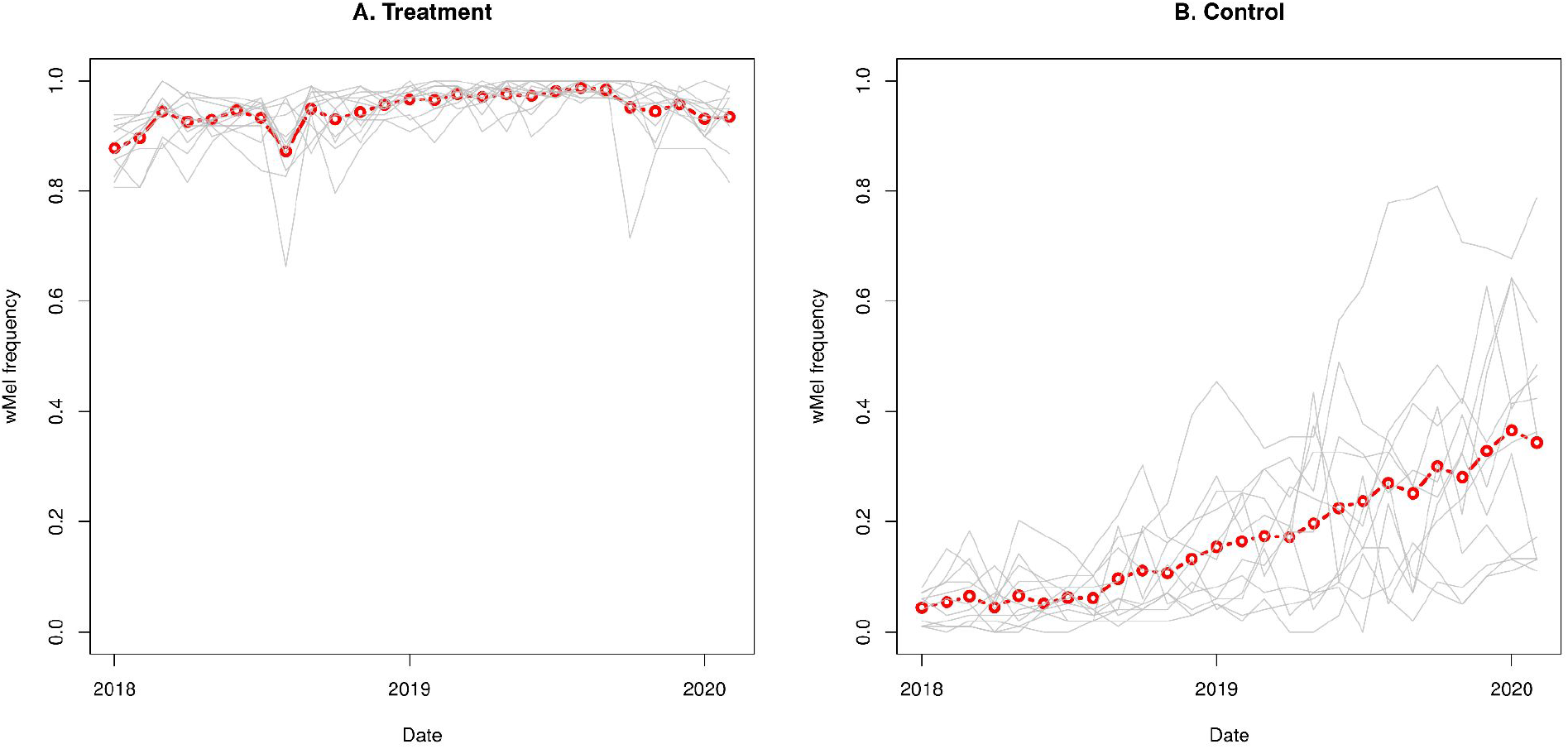
wMel frequency in the AWED trial [7]. Each thin gray line shows the wMel frequency over time in one of the treatment (A) or control (B) clusters. The red line and points show the average of these, which is what was used in the model.

**Fig. S7:**
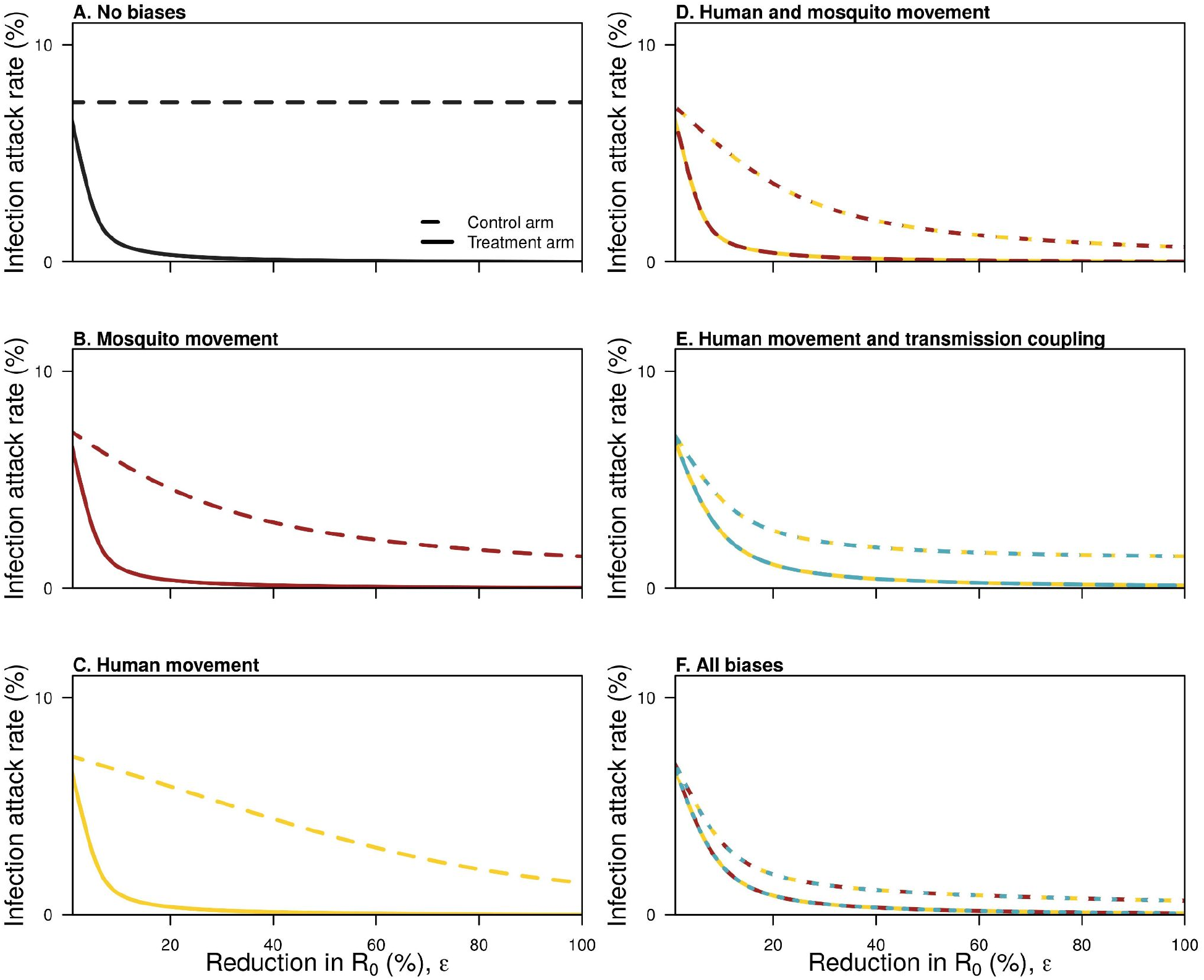
Infection attack rates for each of the six models, delineated by control and intervention arms.

**Fig. S8:**
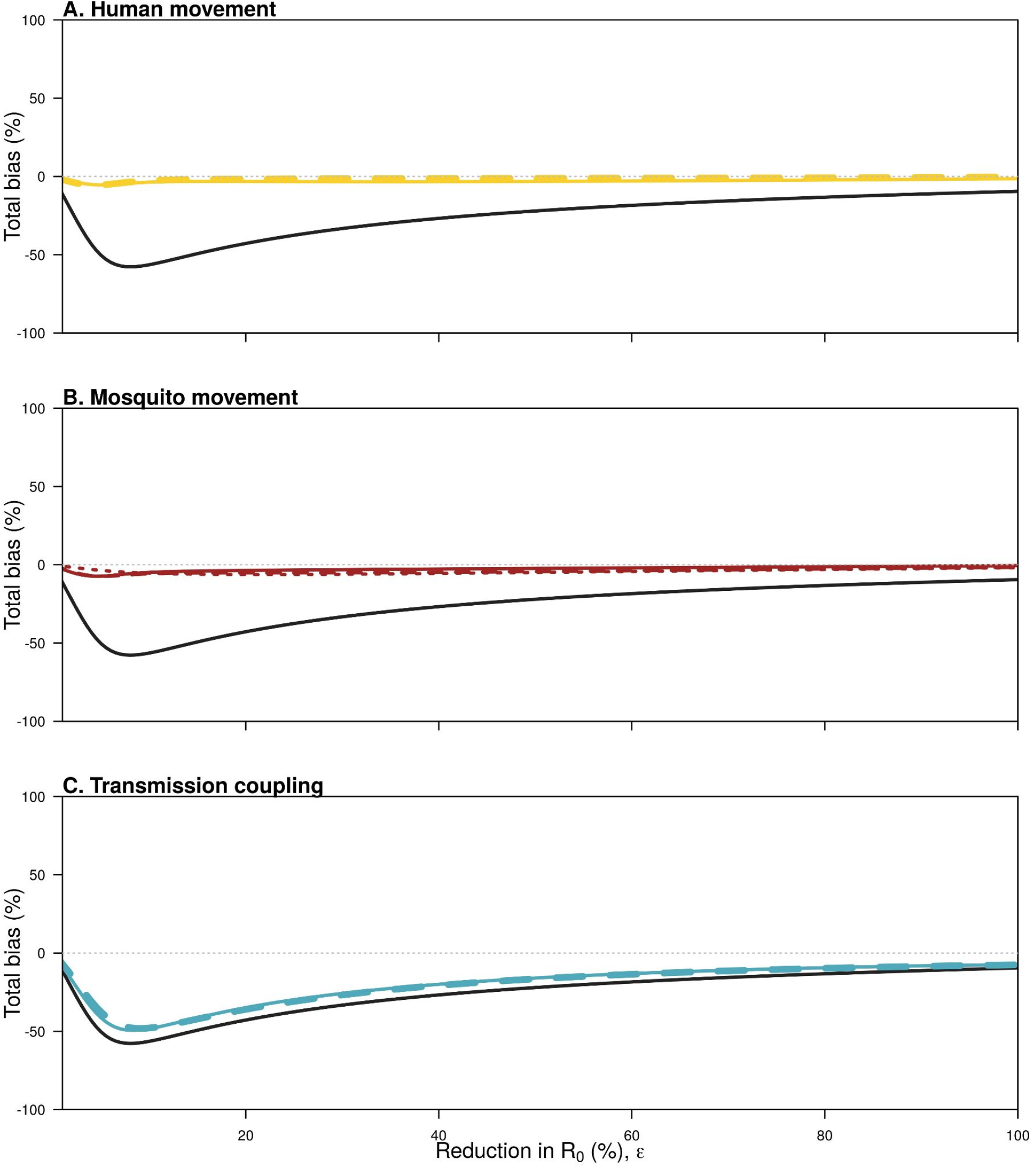
The total bias introduced by each of the three biases. These biases are calculated by subtracting the efficacy of a model with that bias from a model without it, and as biases appear in multiple models there are three possible ways to quantify each bias. These different ways are shown with different line types. For transmission coupling and human movement, two of the ways are equivalent and so these are shown with a thicker line.

**Fig. S9:**
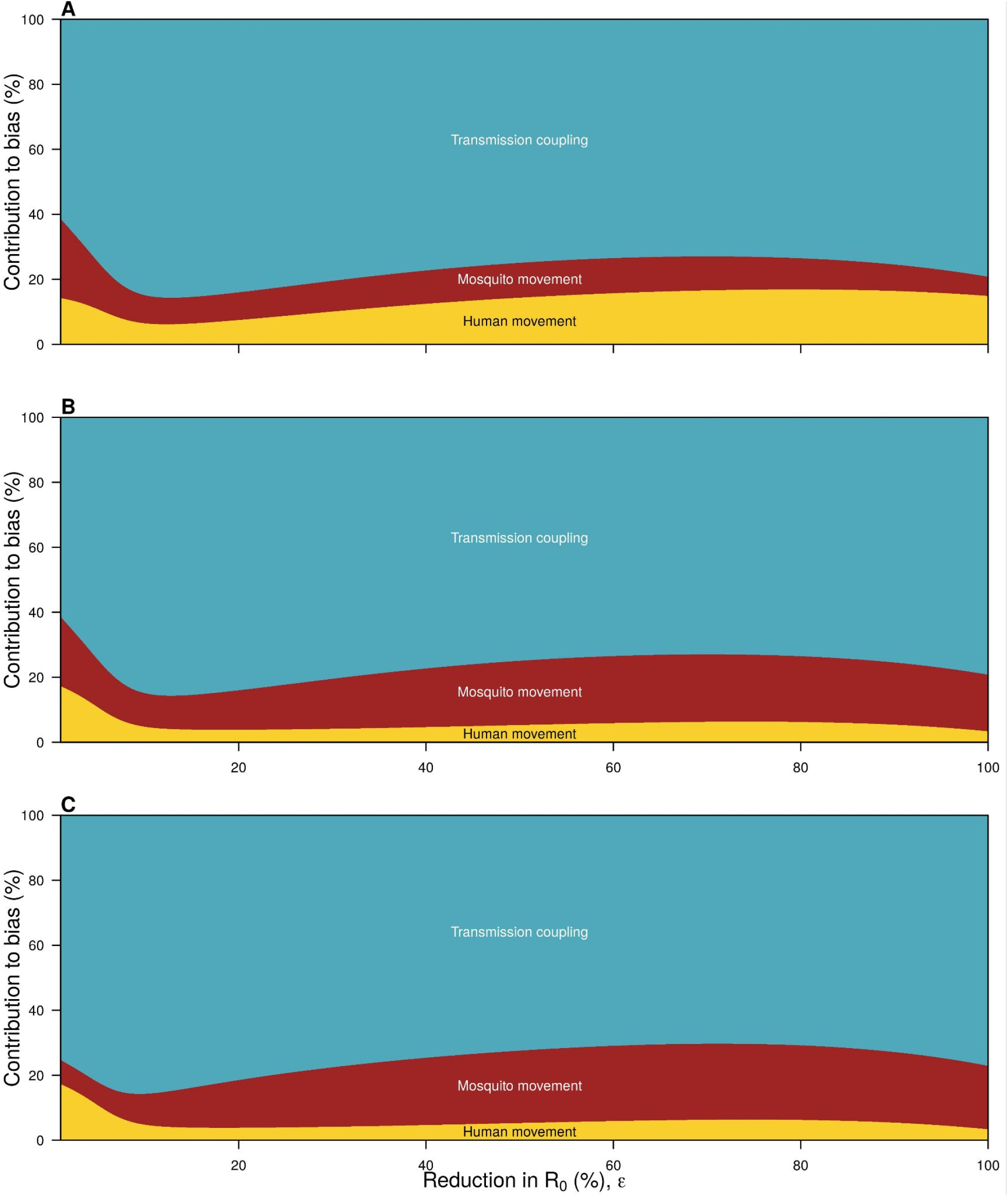
The contribution of each source of bias to the total bias. Each panel shows a different way of calculating the contribution due to that source of bias, which is calculated as the difference in efficacy of a model without that bias and a model with that bias. This can be thought of as embedding the models, and subtracting adjacent pairs of models, so that the sum of each pair of models is equal to the total bias. The embeddings in each panel are: A: no bias→mosquito movement→human movement + mosquito movement→full model. B: no bias→human movement→human movement + mosquito movement→full model. C: no bias→human movement→human movement + transmission coupling→full model.

**Fig. S10:**
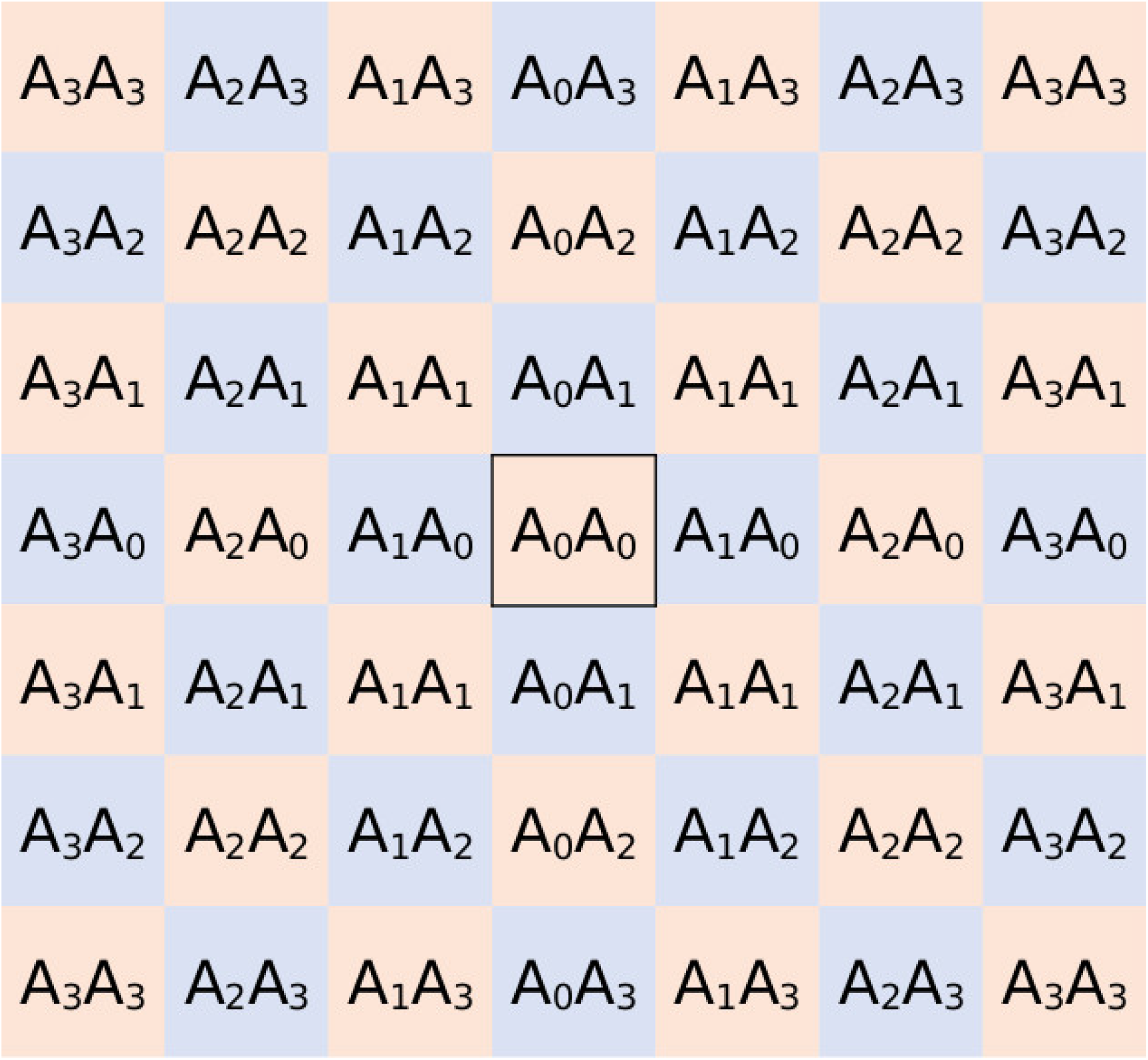
Diagram of the checkerboard arrangement. Cell coloring refers to whether or not someone is the treatment or control cluster. In this case, the central cluster is an individual’s home cluster. A_i_ describes the proportion of time someone spends in a cluster i clusters from their home cluster in one direction. A_i_A_j_ is then the proportion of time someone spends in a cluster i clusters away in one direction, and j in the other.

## References

1. Bhatt S, Gething PW, Brady OJ, Messina JP, Farlow AW, Moyes CL, et al. The global distribution and burden of dengue. Nature. 2013;496: 504–507.

2. Bowman LR, Donegan S, McCall PJ. Is Dengue Vector Control Deficient in Effectiveness or Evidence?: Systematic Review and Meta-analysis. PLoS Negl Trop Dis. 2016;10: e0004551.

3. Morrison AC, Zielinski-Gutierrez E, Scott TW, Rosenberg R. Defining challenges and proposing solutions for control of the virus vector Aedes aegypti. PLoS Med. 2008;5: e68.

4. Ferguson NM, Kien DTH, Clapham H, Aguas R, Trung VT, Chau TNB, et al. Modeling the impact on virus transmission of Wolbachia-mediated blocking of dengue virus infection of Aedes aegypti. Sci Transl Med. 2015;7: 279ra37.

5. Anders KL, Indriani C, Ahmad RA, Tantowijoyo W, Arguni E, Andari B, et al. The AWED trial (Applying Wolbachia to Eliminate Dengue) to assess the efficacy of Wolbachia-infected mosquito deployments to reduce dengue incidence in Yogyakarta, Indonesia: study protocol for a cluster randomised controlled trial. Trials. 2018. doi:10.1186/s13063-018-2670-z

6. Anders KL, Indriani C, Ahmad RA, Tantowijoyo W, Arguni E, Andari B, et al. Update to the AWED (Applying Wolbachia to Eliminate Dengue) trial study protocol: a cluster randomised controlled trial in Yogyakarta, Indonesia. Trials. 2020. doi:10.1186/s13063-020-04367-2

7. Utarini A, Indriani C, Ahmad RA, Tantowijoyo W, Arguni E, Ansari MR, et al. Efficacy of Wolbachia-infected mosquito deployments for the control of dengue. N Engl J Med. 2021;384: 2177–2186.

8. Reiner RC, Achee N, Barrera R, Burkot TR, Chadee DD, Devine GJ, et al. Quantifying the Epidemiological Impact of Vector Control on Dengue. PLOS Neglected Tropical Diseases. 2016. p. e0004588. doi:10.1371/journal.pntd.0004588

9. Stoddard ST, Morrison AC, Vazquez-Prokopec GM, Paz Soldan V, Kochel TJ, Kitron U, et al. The role of human movement in the transmission of vector-borne pathogens. PLoS Negl Trop Dis. 2009;3: e481.

10. Kraemer MUG, Bisanzio D, Reiner RC, Zakar R, Hawkins JB, Freifeld CC, et al. Inferences about spatiotemporal variation in dengue virus transmission are sensitive to assumptions about human mobility: a case study using geolocated tweets from Lahore, Pakistan. EPJ Data Sci. 2018;7: 16.

11. Penny MA, Galactionova K, Tarantino M, Tanner M, Smith TA. The public health impact of malaria vaccine RTS,S in malaria endemic Africa: country-specific predictions using 18 month follow-up Phase III data and simulation models. BMC Med. 2015;13: 170.

12. Penny MA, Verity R, Bever CA, Sauboin C, Galactionova K, Flasche S, et al. Public health impact and cost-effectiveness of the RTS,S/AS01 malaria vaccine: a systematic comparison of predictions from four mathematical models. Lancet. 2016;387: 367–375.

13. Slater HC, Foy BD, Kobylinski K, Chaccour C, Watson OJ, Hellewell J, et al. Ivermectin as a novel complementary malaria control tool to reduce incidence and prevalence: a modelling study. Lancet Infect Dis. 2020;20: 498–508.

14. Achee NL, Gould F, Perkins TA, Reiner RC Jr, Morrison AC, Ritchie SA, et al. A critical assessment of vector control for dengue prevention. PLoS Negl Trop Dis. 2015;9: e0003655.

15. Miller JC. A note on the derivation of epidemic final sizes. Bull Math Biol. 2012;74: 2125–2141.

16. Multerer L, Glass TR, Vanobberghen F, Smith T. Analysis of contamination in cluster randomized trials of malaria interventions. Trials. 2021;22: 613.

17. Multerer L, Vanobberghen F, Glass TR, Hiscox A, Lindsay SW, Takken W, et al. Estimating intervention effectiveness in trials of malaria interventions with contamination. Malar J. 2021;20: 413.

18. Hawley WA, Terlouw DJ, Ter Kuile FO, Gimnig JE, Phillips-Howard PA, Hightower AW, et al. Community-wide effects of permethrin-treated bed nets on child mortality and malaria morbidity in western Kenya. Am J Trop Med Hyg. 2003;68: 121–127.

19. Hayes RJ, Moulton LH. Cluster Randomised Trials. CRC Press; 2017.

20. McCann RS, van den Berg H, Takken W, Chetwynd AG, Giorgi E, Terlouw DJ, et al. Reducing contamination risk in cluster-randomized infectious disease-intervention trials. Int J Epidemiol. 2018;47: 2015–2024.

21. Halloran ME, Auranen K, Baird S, Basta NE, Bellan SE, Brookmeyer R, et al. Simulations for designing and interpreting intervention trials in infectious diseases. BMC Med. 2017;15: 223.

22. Britton T. Stochastic epidemic models: a survey. Math Biosci. 2010;225: 24–35.

23. Cavany SM, Huber JH, Wieler A, Elliott M, Tran QM, España G, et al. Ignoring transmission dynamics leads to underestimation of the impact of a novel intervention against mosquito-borne disease. medRxiv. 2021; 2021.11.19.21266602.

24. Foy BD, Alout H, Seaman JA, Rao S, Magalhaes T, Wade M, et al. Efficacy and risk of harms of repeat ivermectin mass drug administrations for control of malaria (RIMDAMAL): a cluster-randomised trial. Lancet. 2019;393: 1517–1526.

25. James S, Collins FH, Welkhoff PA, Emerson C, Godfray HCJ, Gottlieb M, et al. Pathway to Deployment of Gene Drive Mosquitoes as a Potential Biocontrol Tool for Elimination of Malaria in Sub-Saharan Africa: Recommendations of a Scientific Working Group. Am J Trop Med Hyg. 2018;98: 1–49.

26. United Nations Publications. World Population Prospects 2019: Demographic Profiles. 2020.

27. Burattini MN, Chen M, Chow A, Coutinho FAB, Goh KT, Lopez LF, et al. Modelling the control strategies against dengue in Singapore. Epidemiol Infect. 2008;136: 309–319.

28. Reich NG, Shrestha S, King AA, Rohani P, Lessler J, Kalayanarooj S, et al. Interactions between serotypes of dengue highlight epidemiological impact of cross-immunity. J R Soc Interface. 2013;10: 20130414.

29. Indriani C, Ahmad RA, Wiratama BS, Arguni E, Supriyati E, Tedjo Sasmono R, et al. Baseline Characterization of Dengue Epidemiology in Yogyakarta City, Indonesia, before a Randomized Controlled Trial of Wolbachia for Arboviral Disease Control. The American Journal of Tropical Medicine and Hygiene. 2018. pp. 1299–1307. doi:10.4269/ajtmh.18-0315

30. Kot M, Lewis MA, van den Driessche P. Dispersal data and the spread of invading organisms. Ecology. 1996;77: 2027–2042.

31. Ten Bosch QA, Clapham HE, Lambrechts L, Duong V, Buchy P, Althouse BM, et al. Contributions from the silent majority dominate dengue virus transmission. PLoS Pathog. 2018;14: e1006965.

32. United Nations Publications. World Population Prospects 2019: Demographic Profiles. 2020.

